# Drug interactions in hospital prescriptions in Denmark: Prevalence and associations with adverse outcomes

**DOI:** 10.1101/2021.05.27.21257764

**Authors:** Cristina Leal Rodríguez, Benjamin Skov Kaas-Hansen, Robert Eriksson, Jorge Hernansanz Biel, Kirstine G. Belling, Stig Ejdrup Andersen, Søren Brunak

## Abstract

**Importance:** While the beneficial effects of medications are numerous, drug-drug interactions may lead to adverse drug reactions that are preventable causes of morbidity and mortality.

**Objective:** To quantify the prevalence of potential drug-drug interactions in drug prescriptions at Danish hospitals, estimate the risk of adverse outcomes associated with discouraged drug combinations, and highlight the patient types (defined by the primary diagnosis of the admission) that appear to be more affected.

**Design:** Cross-sectional (descriptive part) and cohort study (adverse outcomes part).

**Setting:** Hospital electronic health records from two Danish regions (approx. 2.5 million people) from January 2008 through June 2016.

**Participants:** Inpatients receiving two or more medications during their admission.

**Exposure:** Concomitant prescriptions of potentially interacting drugs as per the Danish Drug Interaction Database.

**Main outcome and measure:** Descriptive part: prevalence of potential drug-drug interactions in general and discouraged drug pairs in particular during admissions. Adverse outcomes part: post-discharge all-cause mortality rate, readmission rate and length-of-stay.

**Results:** Among 2,886,227 hospital admissions (945,475 patients; median age 62 years [IQR: 41-74]; 54% female; median number of drugs 7 [IQR: 4-11]), patients in 1,836,170 admissions were exposed to at least one potential drug-drug interaction (659,525 patients; median age 65 years [IQR: 49-77]; 54% female; median number of drugs 9 [IQR: 6-13]), and in 27,605 admissions to a discouraged drug pair (18,192 patients; median age 68 years [IQR: 58-77]; female 46%; median number of drugs 16 [IQR: 11-22]). Meropenem-valproic acid (HR: 1.5, 95% CI: 1.1–1.9), domperidone-fluconazole (HR: 2.5, 95% CI: 2.1–3.1), imipramine-terbinafine (HR: 3.8, 95% CI: 1.2–12), agomelatine-ciprofloxacin (HR: 2.6, 95% CI: 1.3–5.5), clarithromycin-quetiapine (HR: 1.7, 95% CI: 1.1–2.7), and piroxicam-warfarin (HR: 3.4, 95% CI: 1–11.4) were associated with elevated mortality. Confidence interval bounds of pairs associated with readmission were close to 1; length-of-stay results were inconclusive.

**Conclusions and Relevance:** Well-described potential drug-drug interactions are still missed and alerts at point of prescription may reduce the risk of harming patients; prescribing clinicians should be alert when using strong inhibitor/inducer drugs (i.e. clarithromycin, valproic acid, terbinafine) and prevalent anticoagulants (i.e. warfarin and NSAIDs) due to their great potential for dangerous interactions. The most prominent CYP isoenzyme involved in mortality and readmission rates was 3A4.

## Introduction

Two drugs are said to interact when the action of one does or may affect the activity, metabolism or toxicity of the other^1^. Drug-drug interactions (DDIs) constitute a particularly important cause of adverse drug reactions (ADRs) as clinical evidence and (when known) their pharmacological mechanisms make them somewhat predictable. Many hospitalised patients take several drugs and polypharmacy^2^ is estimated to affect 40–65% of hospitalised patients^3,4^.

Although the risk of DDIs is proportional to the number of drugs taken^5^, the clinical consequences vary widely, and ADRs rarely occur^6^. Even if uncommon, serious adverse outcomes do cause harm, constitute economic losses and are to some extent preventable. At particularly elevated risk of ADRs are the elderly (often multimorbid and with reduced physiological capacity)^2^ and patients with diseases in organ systems involved in drug metabolism, particularly kidneys and liver^7^. The consequences of DDIs affect both the individual patient and society as a whole: 10–20% of hospital admissions may be attributable to drug-related problems and toxic effects of medication of i.a. DDIs^8,9^, and studies have linked DDIs to prolonged hospitalisation and increased healthcare costs^10–13^.

The electronic medication management systems deployed at public hospitals in Denmark do not systematically flag problematic drug combinations. Even with such systems in place, alert fatigue is a real issue that requires tailoring to optimise their genuine utility^14^. To this end, appropriate evidence about the extent and nature of the problem is needed.

No studies to date have examined the prevalence of potential drug-drug interactions (pDDIs) in hospitals for different patient types and assessed the clinical impact of pDDIs. This study sought to fill this gap and elicit learning points for clinicians to mitigate this issue.

We used electronic health records (EHRs) to (a) elicit the prevalence of discouraged drug pairs and their expected clinical significance and documentation level, (b) identify which patient types are most affected by discouraged pairs, and (c) gauge the association between discouraged pairs and three adverse outcomes: post-discharge mortality, readmission, and length-of-stay (LOS).

## Materials and methods

### Patients and data

We obtained inpatient data for admissions to twelve public hospitals in the Capital Region and Region Zealand, Denmark, from January 2008 through June 2016. The two regions comprise approximately 2.5 million people, about half of the Danish population^15^.

Admissions of individuals using at least two drugs concomitantly were included. We defined concomitant use as temporally overlapping time prescriptions and identified all two-way drug combinations.

Information on admission timing, diagnoses, and medical histories was obtained from the Danish National Patient Register (DNPR)^16,17^, recording data for department-specific visits. DNPR encodes diagnoses with a Danish version of the International Classification of Disease, 10th revision (ICD-10). An admission’s primary diagnoses are recorded retrospectively at discharge. Successive in-hospital visits were combined into admissions if they were at most one day apart.

We marshalled information on dispensed in-hospital drug prescriptions from OPUS-medication (OpusMed) and Electronic Patient Medication (EPM). The latter has been validated^18^ and the former was used in the same manner; both use the WHO Anatomical Therapeutic Chemical (ATC) classification system^19^.

As our pDDI reference we used the Danish Drug Interaction Database (DID), covering predominantly pharmacokinetic interactions based mainly on published results and maintained by specialists in clinical pharmacology under the auspices of the Danish Medicines Agency^20^.

### pDDI prevalence

This descriptive part was cross-sectional. pDDIs were categorised by management recommendation (five levels), clinical significance (five levels), and documentation level (six levels); we only considered the 14,237 (from a total of 18,691) pDDIs with information on all three axes (**Table 1**). The quality of the documentation level is based on the evidence about the significance of the kinetic or dynamic properties.

**Table 1.** Classification of potential drug-drug interactions based on management recommendation, clinical significance, and documentation level published by the Danish Medicines Agency Drug Interaction Database.

| Management recommendation |  |
| --- | --- |
| 1 | The drug combination should always be avoided (discouraged in text). |
| 2 | The drug combination can be used with dose adjustment. |
| 3 | The drug combination can be used with staggered time of ingestion. |
| 4 | The drug combination can be used under certain precautions, i.e. changing the routes of administration. Alternative agents should be considered. |
| 5 | The drug combination can be used. No action needed as the risk of adverse events appears to be small. |
| Clinical significance |  |
| Major | Clinically pronounced/physiological effect with either significant altered therapeutic response or frequent occurrence of serious adverse reactions. |
| Moderate | Clinically moderate/physiological effect with either slightly altered therapeutic response, or rare occurrence of more serious side effects. Serum concentration changes, which in other experiments have been closely associated with the above-mentioned phenomena. |
| Minor | Unchanged or not significantly altered biological response with fewer and easier side effects - or serum concentration changes, which in other studies have not shown significant changes in the biological response. |
| Possible | Pharmacokinetic changes which are not accompanied by known adverse reactions or changes in the biological response. |
| None | Neither kinetic or physiological/clinical changes. |
| Undetermined | Kinetic or physiological/clinical changes that cannot be estimated based on the available documentation. |
| Documentation level |  |
| Well-documented | At least 2 (from different centres) human controlled trials and/or (before and after) trials in relevant individuals with single or multiple steady state trials in the form of either significant kinetic or dynamic changes. |
| Documented | A human controlled study and/or (before and after) study with steady state single or multiple dose trials in the form of either significant kinetic or dynamic changes. |
| Limited documented | Either more than 2 case reports with relevant during and after kinetics or dynamics or human *in vitro* studies with relevant cytochrome P450 (CYP) fractions and concentrations. |
| Poorly documented | 1-2 case reports. Non-conclusive *in vitro* studies. |

Discouraged drug pairs were defined as prevalent when they occurred in more than 10% of admissions of at least one specific patient type, defined as the ICD-10 chapter of the admission’s primary diagnosis. We used standardised difference in proportions to compare imbalances between binary variables, taking an absolute difference above 10% to indicate substantial imbalance^21^.

### Adverse outcomes of exposure to discouraged combinations

In this analytic part of the study, we screened the effect of all discouraged pairs on post-discharge all-cause mortality rate (henceforth, post-discharge mortality), readmission rate and LOS. Only patients’ first admissions were used. We excluded patients whose exposure started outside the hospital for better-defined exposure start. The effects on post-discharge mortality and readmission were estimated with stratified Cox regression models assuming noninformative censoring^22^ and the effects on LOS with stratified Poisson regression models^23^, with exposure to the discouraged drug pair as the sole explanatory variable. We created strata by greedy 1:5 matching on preference score, an extension of the propensity score accounting for target exposure prevalence^24^. The preference score is the probability that a patient be exposed whether this happened or not. Thus, if two patients have (almost) the same preference score but one was exposed and the other not, the exposure is a likely explanation for their difference in outcome^25–27^.

We used Cyclops^28^ to compute high-dimensional propensity scores^26^ with sparse lasso logistic regression models using up to 843 features derived from eight covariates: age at admission (continuous), sex (binary), patient type (one-hot-encoded), diagnoses during admission (ICD-10 level 3, one-hot-encoded), medication burden (continuous), whether the admission was acute or elective (binary), and weighted Elixhauser comorbidity score (Agency for Healthcare Research Quality^29^ version, continuous). Seeking empirical equipoise, outcome models were fit to patients with preference scores between 0.3 and 0.7^24^. The significance level was set to 5%; power analyses were foregone. Estimates with 95% confidence intervals (CI) wider than 100 on the linear scale were omitted.

### Software

We used the R statistical programming language and Python for data processing, analysis, and visualisation. The analysis workflow was built as a Snakemake pipeline^30^ (**eFigure 1)**. The full analytic code is available upon request.

### Ethics

Data were stored and analysed on a secure cloud in Denmark. Registry data access was approved by the Danish Health Data Authority (FSEID-00003092, FSEID-00004491, FSEID-00003724) and the Danish Patient Safety Authority, which at the time was the competent body for approvals regarding research in EHRs, approved journal access and the purpose for the study (3-3013-1731-1).This article observes relevant items in the Strengthening the Reporting of Observational Studies in Epidemiology (STROBE) statement^31^.

## Results

Among the 4,411,576 admissions of 1,481,584 patients identified, we included 2,886,227 admissions (65%) of 945,475 patients (64%) to whom two or more drugs were administered (**eFigure 2**). **Table 2** shows overall and stratified summary statistics for pertinent variables. The 538,620 (57%) women in the cohort contributed 1,551,131 admissions (54%) and 13,122,610 (54%) dispensed prescriptions. Of these, 27,605 admissions (1%) featured discouraged drug pairs and 12,655 (46%) were administrated to women. pDDIs and discouraged drug pairs were observed more frequently in older patients. Further, the median number of prescribed drugs in admissions with discouraged drug pairs (16, IQR: 11-22) was larger than any-pDDI (9, IQR: 6-13) and no-pDDI (4, IQR: 2-6) admissions. Patients exposed to discouraged drug pairs were more ill and had longer admissions and higher in-hospital mortality.

**Table 2.** Overall and stratified summary statistics of included admissions. Values are N (%) and median (interquartile range). pDDI: potential drug-drug interaction. AHRQ: Agency for Healthcare Research Quality.

|  | Overall | No pDDIs | pDDIs | Discouraged drug pairs |
| --- | --- | --- | --- | --- |
| Admissions | 2,886,227 | 1,050,057 (36%) | 1,836,170 (64%) | 27,605 (1%) |
| Women | 1,551,131 (54%) | 565,697 (54%) | 985,434 (54%) | 12,655 (46%) |
| Patients | 945,475 | 553,612 | 659,525 | 18,192 |
| No. prescriptions | 9 (5-15) | 5 (3-8) | 12 (7-19) | 22 (14-36) |
| in women | 8 (4-15) | 4 (3-7) | 12 (7-19) | 22 (14-35) |
| No. unique prescribed drugs | 7 (4-11) | 4 (2-6) | 9 (6-13) | 16 (11-22) |
| Unique prescribed drugs |  |  |  |  |
| 2-4 drugs | 888,934 (31%) | 629,786 (60%) | 259,148 (14%) | 520 (2%) |
| 5-9 drugs | 1,042,023 (36%) | 347,943 (33%) | 694,080 (38%) | 4,408 (16%) |
| ≥ 10 drugs | 955,270 (33%) | 72,328 (7%) | 882,942 (48%) | 22,677 (82%) |
| Age in years | 62 (41- 74) | 51 (30-69) | 65 (49-77) | 68 (58-77) |
| Age group |  |  |  |  |
| < 18 years | 203,125 (7%) | 148,043 (14%) | 55,082 (3%) | 492 (2%) |
| 18-44 years | 619,540 (22%) | 293,005 (28%) | 326,535 (18%) | 2,359 (9%) |
| 45-64 years | 773,558 (27%) | 269,693 (26%) | 503,865 (27%) | 7,790 (28%) |
| 65-74 years | 577,389 (20%) | 162,494 (16%) | 414,895 (23%) | 8,166 (30%) |
| 75-84 years | 461,247 (16%) | 114,094 (11%) | 347,153 (19%) | 6,588 (24%) |
| ≥ 85 years | 251,368 (9%) | 62,728 (6%) | 188,640 (10%) | 2,210 (8%) |
| pDDIs per patient | 1 (0-3) | 0 (0-0) | 2 (1-5) | 9 (5-15) |
| Length of stay in days | 3 (1-6) | 2 (1-4) | 3 (2-7) | 7 (3-15) |
| Acute admission | 2,107,774 (73%) | 765,816 (73%) | 1,341,958 (73%) | 19,746 (72%) |
| In-hospital mortality | 62,830 (2%) | 14,397 (1%) | 48,433 (3%) | 1,252 (5%) |
| Low eGFR (<30ml/min/1.73m<sup>2</sup>) | 109,907 (4%) | 15,198 (1%) | 94,709 (5%) | 2,660 (10%) |
| Elixhauser index (AHQR) |  |  |  |  |
| <0 | 646,561 (22%) | 230,311 (22%) | 416,250 (23%) | 4,893 (18%) |
| 0 | 854,868 (30%) | 399,526 (38%) | 455,342 (25%) | 3,838 (14%) |
| 1-4 | 297,174 (10%) | 104,072 (10%) | 193,102 (11%) | 3,157 (11%) |
| ≥5 | 1,087,624 (40%) | 316,148 (30%) | 771,476 (42%) | 15,717 (57%) |
| Most common drug classes (ATC level 3) |  |  |  |  |
| Other analgesics and antipyretics (N02B) | 1,334,677 (63%) | 501,208 (48%) | 1,334,677 (73%) | 22,024 (80%) |
| Antithrombotic agents (B01A) | 1,038,880 (43%) | 211,944 (20%) | 1,038,880 (57%) | 21,890 (79%) |
| Opioids (N02A) | 917,092 (43%) | 318,180 (30%) | 917,092 (50%) | 18,098 (66%) |
| Anti-inflammatory and antirheumatic products, non-steroids (M01A) | 663,518 (29%) | 178,152 (17%) | 663,518 (36%) | 14,900 (54%) |
| Drugs for peptic ulcer and gastro-oesophageal reflux disease (GORD) (A02B) | 642,650 (27%) | 141,344 (13%) | 642,650 (35%) | 15,429 (56%) |
| Beta-lactam antibacterials, penicillins (J01C) | 686,899 (24%) | 206,305 (20%) | 480,594 (26%) | 10,796 (39%) |
| Loop (high-ceiling) diuretics (C03C) | 518,342 (18%) | 52,487 (5%) | 465,855 (25%) | 131,98 (48%) |
| Most common primary diagnosis |  |  |  |  |
| Abdominal and pelvic pain (R10) | 63,574 (2%) | 26,387 (3%) | 37,187 (2%) | 448 (2%) |
| Pneumonia, organism unspecified (J18) | 60,237 (2%) | 21,369 (2%) | 38,868 (2%) | 1,057 (4%) |
| Atrial fibrillation and flutter (I48) | 55,405 (2%) | 14,422 (1%) | 40,983 (2%) | 667 (2%) |
| Mental and behavioural disorders due to use of alcohol (F10) | 51,347 (2%) | 29,206 (3%) | 22,141 (1%) | 135 (0%) |
| Other chronic obstructive pulmonary disease (J44) | 45,919 (2%) | 16,039 (2%) | 29,880 (2%) | 474 (2%) |
| Nonrheumatic aortic valve disorders (I35) | 10,244 (0%) | 1,784 (0%) | 8,646 (0%) | 1,439 (5%) |
| Acute myocardial infarction (I21) | 30,250 (1%) | 983 (0%) | 29,267 (2%) | 244 (1%) |
| Angina pectoris (I20) | 31,664 (1%) | 5,366 (1%) | 26,298 (1%) | 190 (1%) |
| Bacterial pneumonia, NOC (J15) | 24,045 (1%) | 8,420 (1%) | 15,625 (1%) | 528 (2%) |
| Other sepsis (A41) | 28,401 (1%) | 7,348 (1%) | 21,053 (1%) | 482 (2%) |

Of 344,489 unique drug pairs administered in-hospital, 5,646 (2%) were pDDIs; 1,836,170 admissions (64%) of 659,525 patients (70%) featured at least one of these 5,646 pDDIs. In 27,605 admissions (1%) of 18,192 patients (2%) at least one of the 146 (3%) discouraged drug pairs was used, most with expected major (71%) and moderate (21%) clinical significance (**Table 3** and **eTable 1**). The most prescribed drugs involved in discouraged drug pairs were, in descending order of number of users, pantoprazole (nine admissions [0.0%] of five patients [0.0%] exposed to discouraged drug pairs of 570,440 admissions of 224,002 pantoprazole users), ibuprofen (9,982 admissions [1.8%] of 7,368 patients [2.0%] of 569,223 admissions of 365,302 users), simvastatin (5,048 admissions [1.1%] of 3,887 patients [3.6%] of 442,545 admissions of 148,579 users), metoprolol (1,191 admissions [0.3%] of 399 patients [0.3%] exposed of 379,785 admissions of 127,237 users), and diclofenac (1,917 admissions [1.1%] of 1,326 patients [1.1%] exposed of 177,928 admissions of 120,256 users) affecting up to 3% of the hospitalized patients receiving the drugs (**eTable 2**). In contrast, more uncommon drugs (used by less than 1% of hospitalized patients), e.g. erythromycin (1,573 admissions [27.8%] of 1,461 patients [29.2%] exposed out of 5,665 admissions of 5,001 users), rifabutin (25 admissions [24.8%] of 10 patients [21.7%] exposed out of 101 admissions of 46 users), ketoconazole (644 admissions [20.4%] of 320 patients [21.2%] exposed out of 3,158 admissions of 1,513 users), warfarin (12,570 admissions [10.3%] of 8,791 patients [20.9%] exposed out of 121,653 admissions of 42,101 users), and domperidone (2,872 admissions [12.4%] of 2,028 patients [19.2%] exposed out of 23,213 admissions of 10,571 users) were more often given as part of discouraged pairs (**eTable 2, eFigure 3**).

**Table 3.** Unique drug combinations (upper cells) and prevalence (lower cells) of pDDIs by management recommendation and clinical significance. Values are N (%).

| Recommendation level | Clinical significance |  |  |  |  |  | Total |
| --- | --- | --- | --- | --- | --- | --- | --- |
|  | Major | Moderate | Minor | Possible | None | Un-determined |  |
| 1: Discouraged | 104 (71) | 31 (21) | - | 8 (5) | - | 3 (2) | 146 (3) |
|  | 16,339 (90) | 1,293 (7) | - | 1,206 (7) | - | 24 (0) | 18,192 (3) |
| 2: Dose adjustment | 164 (16) | 457 (45) | 48 (5) | 279 (28) | 1 (0) | 56 (6) | 1,005 (18) |
|  | 40,718 (27) | 91,264 (61) | 12,606 (8) | 58,622 (39) | 25 (0) | 4,953 (3) | 148,455 (23) |
| 3: Staggered ingestion | 53 (23) | 100 (44) | 9 (4) | 47 (21) | - | 17 (8) | 226 (4) |
|  | 14,339 (16) | 38,544 (43) | 244 (0) | 12,264 (14) | - | 43,776 (48) | 90,662 (14) |
| 4: Precautions | 300 (17) | 602 (35) | 86 (5) | 601 (35) | 30 (2) | 123 (7) | 1,742 (31) |
|  | 45,221 (8) | 459,717 (86) | 82,611 (16) | 249,539 (47) | 12,214 (2) | 106,406 (20) | 532,066 (81) |
| 5: No action needed | 6 (0) | 82 (3) | 311 (12) | 206 (8) | 1,648 (65) | 274 (11) | 2,527 (45) |
|  | 165 (0) | 97,285 (21) | 189,797 (40) | 116,185 (25) | 399,935 (85) | 191,767 (41) | 470,956 (71) |
| Total | 627 (11) | 1,272 (23) | 454 (8) | 1,141 (20) | 1,679 (30) | 473 (8) | 5,646 |
|  | 92,167 (14) | 517,599 (78) | 225,512 (34) | 1,679 (0) | 400,935 (61) | 264,711 (40) | 659,525 |

Overall, patients admitted with cardiovascular diseases (ICD-10 chapter IX); endocrine, nutritional and metabolic diseases (chapter IV); and respiratory diseases (chapter X) were more frequently exposed to discouraged drug pairs unlike obstetrical patients (chapter XV) and patients admitted for other reasons (chapter XXI) (**Figure 1A, eFigure 4**). Discouraged pairs varied among the remaining patient types, but within the ±0.1 threshold indicative of negligible imbalance (**Figure 1A**). In contrast, most drugs were more frequently prescribed in admissions with discouraged pairs with many above the 0.1 threshold except misoprostol and oxytocin (**Figure 1B**).

**Figure 1.**
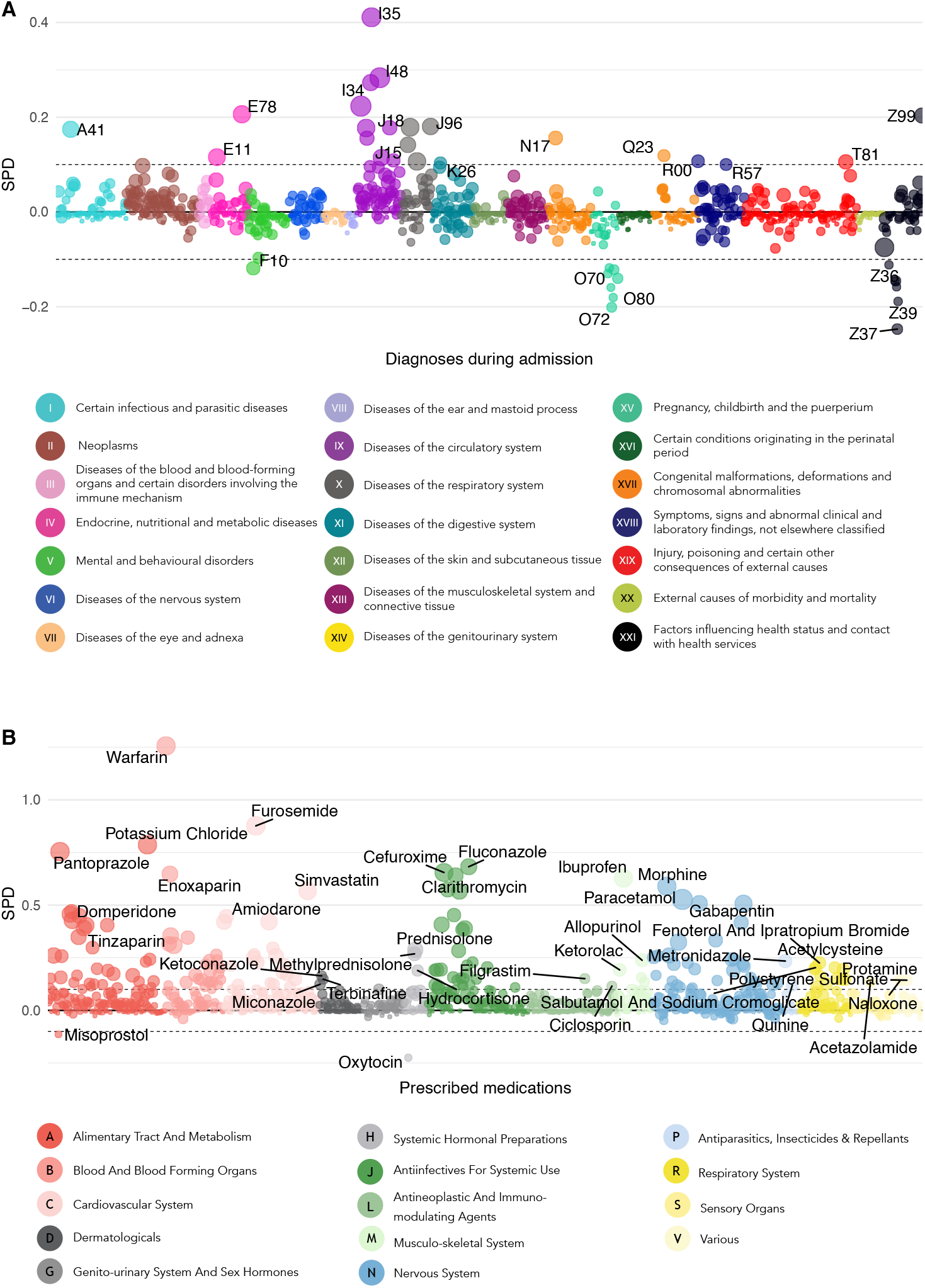
Standardised differences in proportions (i.e. discouraged drug pairs initiated versus not) of diagnoses (panel A) and prescribed drugs during admissions (panel B), respectively. The colour represents ICD-10 chapter and anatomical ATC level, respectively, and the size is the prevalence in patients exposed to discouraged drug pairs. The top three diagnoses and drugs are labelled, and an interactive version of the figure is provided as online supplementary material.

In the 65 discouraged drug pairs (45%) prescribed to five patients or more (**eTable 3**), seven were prevalently (>10% of admissions) prescribed during hospital admissions (**Figure 2**). The most prominent pair was warfarin-ibuprofen, prevalent in all patient types except three (chapters X, XVI and XX). The second-most prominent was simvastatin-clarithromycin, prevalent in six patient types (I, III, IV and X-XII); the third-most was domperidone-fluconazole, prevalent in five patient types (II-IV, VXIII and XXI). The other four were warfarin-diclofenac (XIV, XV, XVII), fluoxetine-venlafaxine (V), meropenem-valproic acid (VI) and erythromycin-fluconazole (XI). **eFigures 4-6** show the prevalence of each drug and each diagnosis in patients exposed vs non-exposed to discouraged drug pairs.

**Figure 2.**
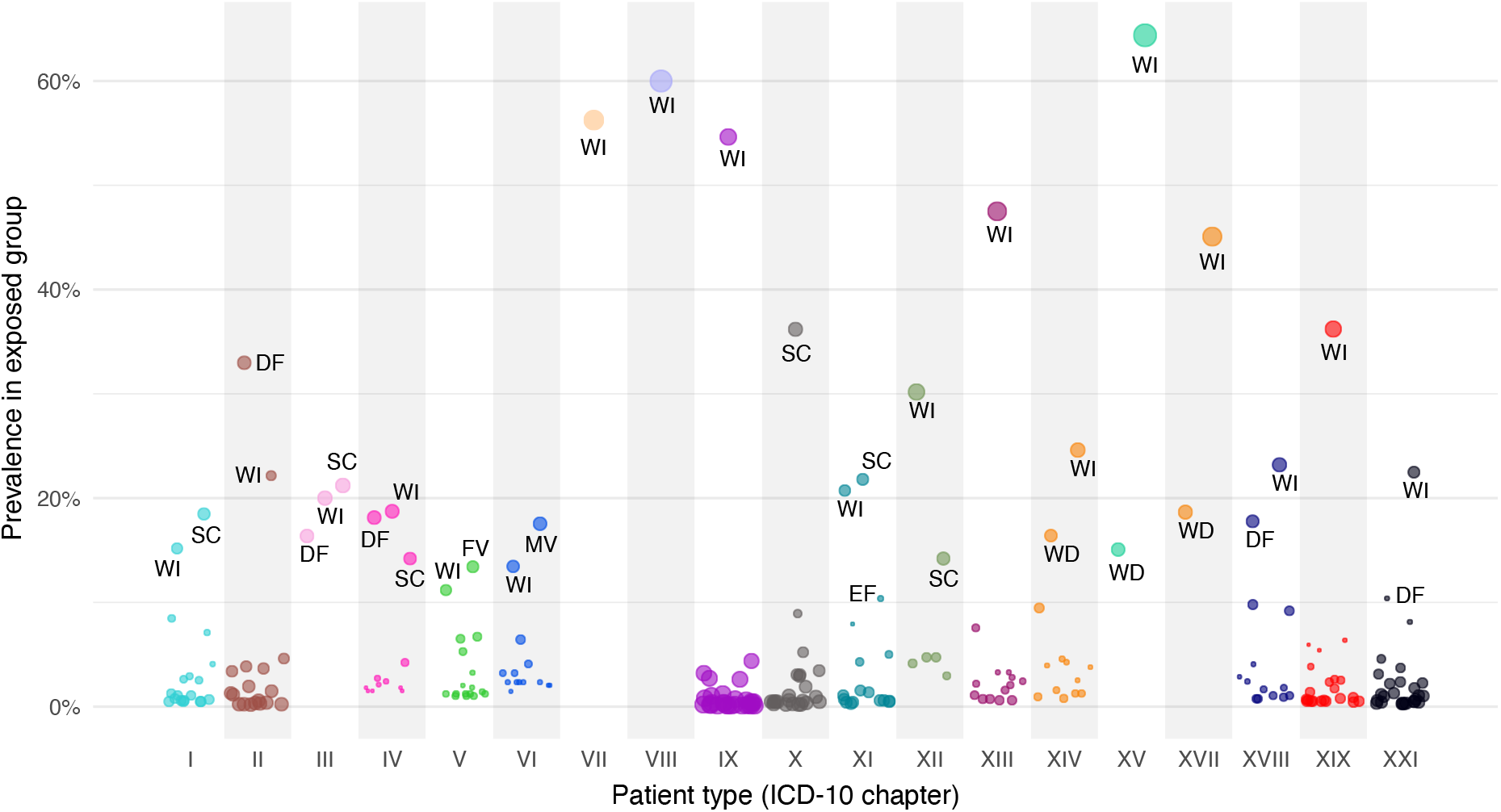
Prevalence of discouraged drug pairs by patient type. Each point represents one discouraged drug pair, and size the absolute value of the standardised difference in proportions using as reference admissions during which treatment with any discouraged pair was initiated. DF (N = 5): Domperidone (A03FA03) + Fluconazole (J02AC01); WD (N = 3): Warfarin (B01AA03) + Diclofenac (M01AB05, systemic); WI (N = 18): Warfarin (B01AA03) + Ibuprofen (M01AE01); SC (N = 6): Simvastatin (C10AA01) + Clarithromycin (J01FA09); MV (N = 1): Meropenem (J01DH02) + Valproic acid (N03AG01); EF (N = 1): Erythromycin (J01FA01) + Fluconazole (J02AC01); FV (N = 1): Fluoxetine (N06AB03) + Venlafaxine (N06AX16).

**Figure 3** shows the estimated effects of exposure on mortality rate, readmission rate and LOS; **eTable 5** contains the numerical estimates. Six discouraged drug pairs were significantly associated with increased mortality rate, of which particularly the 95% CIs of meropenem-valproic acid, domperidone-fluconazole, imipramine-terbinafine and agomelatine-ciprofloxacin are relatively far from 1. Ertapenem-fluconazole, amitriptyline-terbinafine as well as clarithromycin with ticagrelor, tacrolimus and everolimus, respectively, were associated with substantially elevated readmission rates albeit with CI bounds near 1.

**Figure 3.**
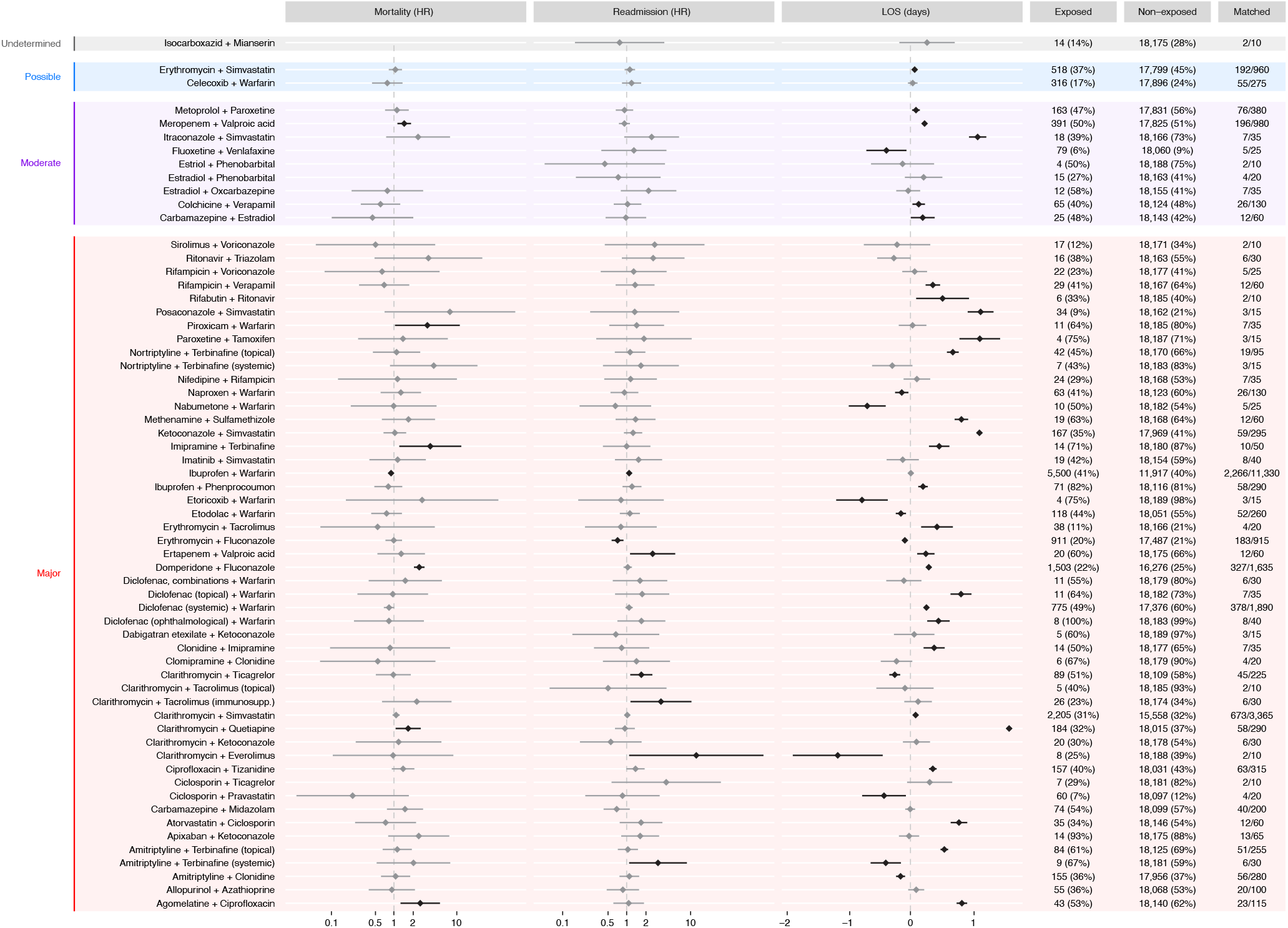
Estimate effect sizes of exposure to discouraged drug pairs and post-discharge mortality rate (hazard ratio, HR), readmission rate (HR) and length-of-stay (change in days). Diamonds show point estimates of the effect sizes, horisontal lines the 95% confidence intervals. The exposed and non-exposed columns show count (empirical equipoise) and the matched column shows the number of exposed/non-exposed used to estimate the effects of that pair.

Many discouraged pairs were associated with longer or shorter hospital stays with most effect sizes within approximately ±1 day.

## Discussion

We found that 1,836,170 admissions (64%) of 659,525 patients (70%) featured at least one pDDI and that during 27,605 admissions (1%) of 18,192 patients (2%) at least one discouraged drug pair was administered. Seven discouraged pairs were prevalent, most notably warfarin-ibuprofen (18 patient types), simvastatin-clarithromycin (six patient types) and domperidone-fluconazole (five patient types). Of the prevalent discouraged pairs, domperidone-fluconazole and meropenem-valproic acid (one patient type) were significantly associated with elevated mortality. The prevalent pair warfarin-ibuprofen was just statistically significantly associatiated with elevated readmission rates and three of five discouraged pairs associated with elevated readmission rates involved clarithromycin. LOS results were inconclusive.

The increasing availability of longitudinal patient data and growing access to databases with DDI information facilitate comparative, data-driven approaches to identify, anticipate and explain DDIs^32^. Indeed, in this study we used comprehensive phenotypic in-hospital data to detail the landscape of in-hospital pDDIs with particular focus on discouraged drug pairs to elicit their effects on potentially preventable adverse outcomes.

Prevalence patterns and effects of pDDIs are elusive because many potentially interacting drug combinations offer genuine clinical utility if used consciously by alert physicians.

Teasing apart these dynamics is difficult on a large scale. Indeed, studies of pDDI prevalence in hospitalised patients often use relatively small samples from sub-populations such as critically ill or oncological patients^33–35^. Our approach was different seeking to conduct a large-scale screening of hospitalised patients, focusing on outright discouraged drug pairs because their clinical benefits unlikely outweigh their potential harm.

A recent systematic review of clinically manifested DDIs^36^ found prevalence estimates up to 26% in not-critically-ill hospitalised patients^37^. Further, the number of drugs used concomitantly has been shown to be a significant risk factor for interactions at hospitals and in primary care^38–41^. We also observed widespread polypharmacy among patients exposed to pDDIs especially when exposed to discouraged drug pairs. However, unlike for diagnoses, no drugs involved in pDDIs emerged as neither particularly frequent nor infrequent except misoprostol and oxytocin. Thus, perceiving the effect of polypharmacy solely in terms of the association between number of concomitant drugs and pDDIs is arguably of limited use as it tells us little about the nature of the association. Instead, other phenotypic factors such as comorbidities may be of greater utility to the prescribing physician at point of care.

A Danish study of 167,232 patients from 1998 on the island of Funen found that 4.4% of all inhabitants of age above 70 were prescribed drug combinations with a high risk of severe interactions^42^. A recent Brazilian study with approximately 340,000 patients from primary- and secondary-care hospitals arrived at a similar figure^43^. These estimates are substantially lower than our 14% patients prescribed pDDIs with expected major clinical significance (**Table 3**), likely because our data are newer than those in the former and include also tertiary hospitals unlike both those studies.

Another study from Denmark published in 2005 found that pDDIs are prevalent but mostly clinically insignificant^44^. Our results agree with this notion: six discouraged combinations featured substantial and statistically significant associations with elevated mortality, of which only two were prevalent in particular patient types (meropenem-valproic acid, domperidone-fluconazole). This was the case for only warfarin-ibuprofen with respect to readmission rates.

Rarely used drugs are more often involved in potentially dangerous DDIs perhaps due to prescribers’ lack of specific knowledge on these drugs; consider three examples. First, using meropenem (or ertapenem) with valproic acid elevates the risk of seizures (unknown mechanism) and meropenem consumption is increasingly prescribed at emergency departments, often by junior doctors. Second, the cardiac risks of domperidone and erythromycin (prolonged QT and Torsades-de-Pointes) are aggravated by concurrent use of fluconazole (or any conazole) because the latter impedes their metabolism by inhibiting CYP3A4^45^. Third, concurrent use of agomelatine and ciprofloxacin increases the exposure of the former because the later inhibits CYP1A2. Causes of death in deceased exposed to these drug pairs did not suggest unexpected patterns (**eFigure 7**).

Some active ingredients involved in discouraged drug pairs have several ATC codes (terbinafine, diclofenac and tacrolimus) and the somewhat agreeing effect estimates on mortality and readmission rates add confidence to these findings. Interestingly, the effects on LOS were not consistent across ATC codes for the same active ingredient prompting cautious interpretation. Indeed, LOS is elusive: for example, a short admission can end with discharge to home or death. To arrive at meaningful conclusions on lengths-of-stays, one would need to use for example drug administrations allowing for time-to-event analyses, something not possible with these data.

## Strengths and limitations

This study features a range of strengths. First, this is the largest study assessing the prevalence of discouraged drug pairs and their effects on adverse outcomes among hospitalised patients. Second, we used unfiltered data from a heterogeneous population of almost one million hospitalised patients over an eight-year period. Third, detailed and reliable register data allow for detailed phenotyping, both with respect to diagnoses and medication use. Fourth, such deep phenotyping underpins the use of high-dimensional preference scores to obtain approximate empirical equipoise when studying the associations between exposure and adverse outcomes. Fifth, the risk of selection bias and loss to follow-up was minimal.

Nonetheless, there are potential weaknesses. First, hospital data may be subject to recall and information bias. This is likely not an issue for this study because we rely on near-objective data (e.g. validated source of medication data) used also for administrative and billing purposes. Bias by indication could be a problem but the use of high-dimensional propensity scores should, at least in part, counter this. Second, we only considered two-way pDDIs.

Large-scale screening for N-drug interactions is difficult due to combinatorial explosion in the number of possibilities and difficulties in defining a proper reference to which the results should be compared. Instead, targeted investigations would be meaningful, e.g. on triple whammy and its effect on kidney function. Third, different pDDI databases likely feature discrepancies regarding management recommendations and clinical significance, and the DID covers primarily pharmacokinetic interactions. Further, DID allows different levels of evidence: for older drugs only pDDIs supported by published evidence are considered, whereas for newer drugs also pDDIs from summaries of product characteristics not published elsewhere are included. This database, nonetheless, is well-known among Danish physicians and used in daily practice to guide medicinal treatment and, as such, makes for a natural gold standard against which to compare real-life prescriptions in Denmark. Fourth, the pDDIs involving antibiotics and systemic antifungals and associated with elevated mortality are used to treat serious infections. Thus, exposure to these combinations could be proxies for serious clinical conditions, themselves associated with high mortality. If so, physicians could have deemed it worthwhile to use a discouraged drug pair due to bleak prognoses. Fifth, despite a large dataset we had relatively few patients exposed to several discouraged drug pairs, making it difficult to rule out effects of these exposures on mortality and readmission rates even though we did not find any.

## Conclusion

Discouraged drug pairs are common in hospitalised patients at large and so are potentially problematic drug pairs, notably, combinations of warfarin and NSAIDs and with antiinfectives (especially, azoles, carbapenems and macrolides). The meropenem-valproic acid and domperidone-fluconazole combination, both prevalent in at least one patient type, were significantly associated with elevated post-discharge mortality rate. This study elicited unfortunate prescription patterns with potentially detrimental effects in hospitalised patients and the CYP3A4 isoenzyme was involved in more than half the discouraged pairs associated with elevated mortality or readmission rates.

## Data Availability

Permission to access and analyse the underlying person-sensitive data can be obtained following approval from the Danish Data Protection Agency and the appropriate Danish Region (regional office for registry research in patient journals).

## Competing interests

S.B. reports ownerships in Intomics A/S, Hoba Therapeutics Aps, Novo Nordisk A/S, Lundbeck A/S and managing board memberships in Intomics A/S outside the submitted work. All other authors report no competing interests.

## Acknowledgements

R.E. proposed the idea. C.L.R. and B.S.K.H. designed and conducted the study. S.B. and S.E.A. co-created the project that made available and curated the EHR data. S.B. and S.E.A. obtained funding. C.L.R., B.S.K.H. and J.H.B. performed pre-processing of the data. (B.S.K.H. extracted eGFR measurements, and calculated comorbidity scores; J.H.B. extracted the information from Danish interaction database under the supervision of C.L.R.

C.L.R pre-processed the clinical and medication data, calculated the treatment exposures and treatment overlaps). C.L.R. and B.S.K.H. performed the computational and statistical analysis. B.S.K.H. did the adverse outcome modelling. C.L.R. and B.S.K.H. interpreted data and provided critical intellectual content, under guidance by S.E.A. C.L.R. performed the literature search. C.L.R. and B.S.K.H. wrote the initial draft. All authors have contributed to and approved the final manuscript.

This work was supported by the Novo Nordisk Foundation (grants NNF14CC0001 and NNF17OC0027594) and the Danish Innovation Fund (grant 5153-00002B). The funding bodies had no role in the design and conduct of the study; collection, management, analysis, and interpretation of the data; preparation, review, or approval of the manuscript; or the decision to submit the manuscript for publication. C.L.R and B.S.K.H. had full access to all the data in the study and takes responsibility for the integrity of the data and the accuracy of the data analysis.

## Notes

### Clinical Trial

Not applicable

### Author Declarations

Data were stored and analysed on a secure cloud in Denmark. Registry data access was approved by the Danish Health Data Authority (FSEID-00003092, FSEID-00004491, FSEID-00003724) and the Danish Patient Safety Authority, which at the time was the competent body for approvals regarding research in EHRs, approved journal access and the purpose for the study (3-3013-1731-1).

