## Supplement for "Drug interactions in hospital prescriptions in Denmark: Prevalence and associations with adverse outcomes": CLR_BSKH_supplement.html

#### Cristina Leal Rodríguez & Benjamin Skov Kaas-Hansen

#### 23 December 2020

- Tables
  - eTable 1: Prevalence of potential drug-drug interactions based on recommendation and documentation level.
  - eTable 2: Drugs involved in discouraged drug pairs.
  - eTable 3: Overview of drugs involved in discouraged combinations.
  - eTable 4: List of discouraged drug pairs.
  - eTable 5: Effect-size estimates of exposure to discouraged drug pairs on post-discharge mortality, readmission and length-of-stay.
- Figures
  - Figure 1A (interactive): Differences in diagnoses between patients exposed and not exposed to discouraged drug pairs.
  - Figure 1B (interactive): Differences in medication exposure between patients exposed and not exposed to discouraged drug pairs.
  - Figure 2 (interactive): Prevalence of discouraged drug pair by patient type.
  - eFigure 1: Pipeline workflow.
  - eFigure 2: Attrition diagram.
  - eFigure 3: Drugs prevalently involved in discouraged drug pairs.
  - eFigure 4: Standardised differences in proportions of diagnoses in admissions with and without discouraged drug pairs by patient type.
  - eFigure 5: Phenotyping of diagnoses and medications.
  - eFigure 6: Phenotyping of diagnoses and medications by patient type.
  - eFigure 7: Causes of death at ICD-10 chapter level among deceased exposed to selected drug pairs.

---

This document contains all supplementary tables and figures for the above-mentioned paper, as well as an interactive version of figure 1 from the main text.

---

### Tables

#### eTable 1: Prevalence of potential drug-drug interactions based on recommendation and documentation level

Unique drug combinations (upper cells) and prevalence (lower cells) of pDDIs by management recommendation and documentation level. Values are N (%).

#### eTable 2: Drugs involved in discouraged drug pairs

Drugs used by less than 5 patients were omitted to avoid privacy issues. The total numbers of admissions and patients are 2,886,227 and 945,475 (denominators in the "overall (%)" columns).

#### eTable 3: Overview of drugs involved in discouraged combinations

Drugs used by less than 5 patients were omitted to avoid privacy issues.

#### eTable 4: List of discouraged drug pairs

Discouraged drug pairs dispensed in at least 5 admissions (≥5 patients) prescribed during patient hospitalisation.

#### eTable 5: Effect-size estimates of exposure to discouraged drug pairs on post-discharge mortality, readmission and length-of-stay

Provides numerical estimate illustrated in Figure 3: estimate (95 confidence interval). Mortality and readmission estimates are hazard ratios. Values below 1 are shown with 2 significant digits, otherwise with 1 significant digit.

### Figures

#### Figure 1A (interactive): Differences in diagnoses between patients exposed and not exposed to discouraged drug pairs.

Diagnoses assigned to between 1 through 4 exposed patients are redacted.

#### Figure 1B (interactive): Differences in medication exposure between patients exposed and not exposed to discouraged drug pairs.

Drug dispensed to between 1 through 4 exposed patients are redacted.

#### Figure 2 (interactive): Prevalence of discouraged drug pair by patient type.

#### eFigure 1: Pipeline workflow

Snakemake pipeline depicting the workflow used for the adverse outcomes analytic part of the study. Schematic overview of the analytic pipeline. Each box represents a distinct step, and arrows represent dependencies between tasks. Preprocessing not depicted.

#### eFigure 2: Attrition diagram

#### eFigure 3: Drugs prevalently involved in discouraged drug pairs

Drugs prevalently involved in discouraged drug pairs by management recommendation (columns) and clinical significance (rows), defined as drugs involved in discouraged drug pairs occurring in at least 10 of admissions. Points represent pDDIs, size the drug prevalence, and colour the clinical significance.

#### eFigure 4: Standardised differences in proportions of diagnoses in admissions with and without discouraged drug pairs by patient type

Standardised differences in proportions (i.e. discouraged drug pairs initiated versus not) of diagnoses and during admissions. The colour represents ICD-10 chapter, and the size is the prevalence in patients exposed to discouraged drug pairs. The top three diagnoses and drugs are labelled.

#### eFigure 5: Phenotyping of diagnoses and medications

Detailed diagnoses and medication phenotyping of patients exposed to discouraged drug pairs vs. those not exposed. The plot compares the diagnoses and medications during hospitalisation. Each dot represents one of these covariates with the colour indicating the absolute value of the standardized difference in proportion (SPD). The top three medications and diagnoses with SPD >0.1 (exposed; red) are highlighted.

#### eFigure 6: Phenotyping of diagnoses and medications by patient type

Same as eFigure 5 but by patient type.

#### eFigure 7: Causes of death at ICD-10 chapter level among deceased exposed to selected drug pairs
